# ChatGPT Is Still Not Good Enough at Giving Care-Seeking Advice, or Is It?

**DOI:** 10.1101/2025.05.13.25327519

**Authors:** Marvin Kopka, Longqi He, Markus A. Feufel

## Abstract

Artificial Intelligence tools like ChatGPT are increasingly used by patients to support their care-seeking decisions, although the accuracy of newer models remains unclear. We evaluated 16 ChatGPT models using 45 validated vignettes, each prompted ten times (7,200 total assessments). Each model classified the vignettes as requiring emergency care, non-emergency care, or self-care. We evaluated accuracy against each case’s gold standard solution, examined the variability across trials, and tested algorithms to aggregate multiple recommendations to improve accuracy. o1-mini achieved the highest accuracy (78%), but we could not observe an overall improvement with newer models – although reasoning models (e.g., o4-mini) improved their accuracy in identifying self-care cases. Selecting the lowest urgency level across multiple trials improved accuracy by 4 percentage points. Although newer models slightly outperform laypeople, their accuracy remains insufficient for standalone use. However, making use of output variability with aggregation algorithms can improve the performance of these models.

## Introduction

Artificial Intelligence (AI) applications are increasingly used in healthcare for tasks such as diagnosing patients, summarizing information, and answering health-related questions.^1^ Although AI has traditionally been developed for such specific use cases, general-purpose foundation large language models (LLMs) – such as OpenAI’s GPT or Anthropic’s Claude models – have received increasing attention because they perform well across various domains and tasks.^1^ For example, GPT-4 has passed several medical licensing exams, has proven effective in drafting letters and clinical notes, and appears to be useful in generating differential diagnosis lists with relatively high accuracy.^2–10^ ChatGPT and similar large language models, however, are increasingly used by patients, because they are easy to access, easy to use, and applicable for a variety of different use cases.^11–13^ For example, they are used for answering general health-related questions and for diagnosing symptoms – both use cases in which ChatGPT has demonstrated moderate to high accuracy.^12,14–16^ In terms of answering health-related questions, ChatGPT has been found not only to achieve accuracy levels comparable to medical experts but also to be perceived as more empathetic than medical professionals.^17,18^ For self-diagnosis use cases, it has shown moderate accuracy compared to other self-diagnosis application.^7^ Given that self-diagnosis is not particularly useful for laypeople,^19^ using ChatGPT for obtaining care-seeking advice (which is also referred to as ‘self-triage’, ‘triage advice’, ‘urgency advice’ or ‘dispositional advice’) is said to be a more relevant use case for patients.^19^

Applications for care-seeking advice, which are not based on LLMs, have been extensively tested and yielded mixed results. Some applications outperform medical laypeople and approach the performance of medical professionals, whereas others perform worse than chance.^20–22^ To date, few studies have benchmarked LLMs for care-seeking advice. Among those that have, most report a medium overall accuracy of about 70%, and a performance generally lower than that of medical professionals.^7,16,23,24^ However, most of these studies have examined only a single LLM. Recent benchmarking tests from other domains suggest that newer models, such as GPT-4o and chain-of-thought models (that simulate human meta-cognition by critically assessing and revising their answer before giving output) like o1, perform better than their predecessors.^10,25^ This suggests that ChatGPT models may also be improving their ability to provide care-seeking advice. However, no prior research has explicitly tested this hypothesis.

In addition to overall accuracy, also output variability of LLMs across multiple trials should be examined. Although output variability has been identified as a challenge when LLMs were used by professionals for diagnosing and treating patients,^6–10^ it has not yet been examined in the context of care-seeking advice. If output variability exists, it may limit the interpretability of single-trial LLM evaluations. However, such variability could also be beneficial: research on ‘the wisdom of the crowd’ has shown that aggregating advice from multiple human advisors can improve the accuracy of decisions, and a similar approach may also improve the advice quality of LLMs.^26^ To address these gaps, this study reports a benchmarking test with a longitudinal analysis to (a) compare the accuracy of care-seeking advice across all available ChatGPT models – as the most widely used LLM family^27^ –, (b) analyze the variability of care-seeking advice across multiple trials, and (c) determine whether aggregating advice for the same case across multiple trials can be used to improve accuracy.

## Methods

### Study Design

This study was designed as a longitudinal observational study comparing all available ChatGPT models. The primary outcome is the overall accuracy of the model’s care-seeking advice. As secondary outcomes, we included additional accuracy metrics that have been proposed for evaluating applications for care-seeking advice in previous studies: the accuracy across different urgency levels (emergencies, non-emergencies, and self-care cases) and over- and undertriage errors (higher and lower urgency than appropriate). Also, we assessed the variability of the provided advice.^28,29^ To determine how advice variability may be utilized to improve the accuracy of the model output, we also applied several pooling algorithms to aggregate the advice of the same model across multiple trials and assessed how they impact the accuracy measures identified above.

### Models

To capture trends in accuracy over time, all models released by OpenAI were eligible for inclusion. These models were identified through a document search of OpenAI’s API documentation. The identified models can be categorized into two groups: generative pre-trained transformer (GPT) models and chain-of-thought (CoT) models. CoT models are built on top of GPT models and are automatically instructed to reason through an answer before generating a response. As a result, they take longer to produce an answer but are expected to improve response quality. Because each new model is expected to outperform its predecessors, we sorted all included models by release date.

### Case Vignettes

We used a set of use-case specific case vignettes to evaluate the models, which was developed and validated in previous studies.^24,30^ The set consists of real patient cases taken from individuals who described their symptoms in an online community to receive guidance and information on whether and where to seek care with their problems or symptoms. In total, 45 case vignettes were stratified in the set to reflect the natural base rate of both (1) emergency, non-emergency and self-care cases among patients using the internet for care-seeking advice, and (2) their symptom types according to the Centers for Disease Control and Prevention National Ambulatory Medical Care Survey coding protocol.^24,31^ The chosen case vignette set thus has high external validity for generalizing to real-world scenarios in which medical laypeople use online decision support to determine whether and where to seek care for their health problems. However, given the naturally low frequency of emergencies, the set includes only two case vignettes describing emergency cases. The gold standard solutions for the cases were determined by two licensed physicians who independently rated the cases. In cases of disagreement, they discussed the case until reaching a consensus.^24^

### Outcome Evaluation

To assess the accuracy of care-seeking advice, each model was asked to classify each case vignette and assign the most appropriate triage urgency level. Following established evaluation methods for symptom checkers, the models were required to choose between three urgency options: emergency care, non-emergency care (including a visit to the family physician), or self-care (i.e., watchful waiting and not consulting a healthcare professional).^28,32^ To minimize input variability and potential bias and to increase comparability across studies, we used a standardized one-shot prompt to obtain responses. The prompt was developed in a previous study, which itself was adapted from two earlier studies.^16,23,24^ The prompt reads:

“Please give a triage classification for the following vignette. The criteria for triage classifications are presented below: Emergency: Call 911 or go directly to the emergency room; Non-Emergency: Try to see a doctor, but the symptoms do not require immediate attention at an emergency room.

For example, you can see a doctor in the next week. Self-care: Let the health issue get better on its own and review the situation in a few days again. [Case Vignette]”.

To examine variability in advice, this prompt was used 10 times for each case description.^10^ Model recommendations were obtained via the OpenRouter API using a custom-built Python script. This script added each case description to the prompt and then sent it to the OpenRouter API. To minimize bias, the context window was cleared before each API call, and the next case vignette was processed separately. The API requests were made from Berlin, Germany, on January 9, 2025. Because new models were released after the initial data collection, we conducted two additional rounds of API calls for these models on February 13, 2025 (for o3-mini and o3-mini-high) and on April 23, 2025 (for o3, gpt-4.1-nano, gpt-4.1-mini, gpt-4.1, gpt-4.5-preview, o4-mini and o4-mini-high). The corresponding advice – which included the urgency level and background information or reasons for selecting this urgency level – was recorded and classified into ‘emergency care’, ‘non-emergency care’, or ‘self-care’ in a sequential procedure: first, two researchers manually classified a random selection of 50% of all cases. In cases of disagreement, both researchers re-evaluated the answer provided for the case and discussed it to determine the final classification. In a second step, we employed natural language processing (NLP) to automatically classify the models’ output using gpt-4.1-nano and gpt-4.1-mini as cost-efficient but highly accurate models, each of which classified all outputs separately using a zero-shot prompt. We then assessed any disagreements between these two models manually and compared their classification performance with the manual coding of all cases as the ground truth. Because this procedure yielded satisfying results, the other 50% of all cases were automatically classified using NLP and manual classification in case of disagreements between the two models.

The advice was classified as either correct (if it matched the gold standard solution) or incorrect (if it deviated from the gold standard solution). The primary outcome – overall accuracy – was calculated as the proportion of correctly classified cases across all repeated trials and urgency levels. Secondary outcomes were defined as follows: accuracy for each urgency level was calculated as the proportion of correctly classified cases within that specific urgency level. Overtriage errors were defined as cases in which the model’s recommendation was of higher urgency than the gold standard solution, whereas undertriage errors were defined as cases in which the recommendation was of lower urgency then the gold standard solution. To assess each model’s variability, Cohen’s Kappa was calculated as a measure of agreement across ten trials for the same case. Additionally, to classify different levels of variability, advice for each model was categorized as follows: ‘always correct’ if every trial for a case yielded the correct result, ‘sometimes correct’ if some but not all trials contained the gold standard solution, or as ‘never correct’ if no single trial contained the gold standard solution.

To determine whether accuracy can be improved when using LLMs by aggregating multiple trials, we pooled the recommendations of all trials for each case and model and recalculated the accuracy (and accuracy across each urgency level). We applied four pooling algorithms: the first approach determined whether at least one of the 10 trials contained the gold standard solution. The second algorithm applied the majority rule by selecting the most common recommendation across all trials. The third algorithm used the highest urgency level that appeared in all trials for a case to overcome undertriage errors. The fourth pooling approach used the lowest urgency included in all trials for a case to overcome overtriage errors.

### Data Analysis

The data were analyzed in R using the symptomcheckR, DescTools, and tidyverse packages.^29,33,34^ First, we calculated a two-way mixed, agreement, average-measures intra-class correlation (ICC) to assess inter-rater reliability between the two coders. We then determined the average accuracy and accuracy for each urgency level by calculating the proportion of correct responses. Hence, we treated correctness as a binary variable. Next, we used confusion matrices to visualize over- and undertriage errors, i.e., to assess the direction of disagreement between the models’ recommendations and the gold standard solution. In this analysis, we treated the model’s recommendations as an ordinal variable. We then calculated Cohen’s Kappa for each model to assess the degree of variability. Values below .40 were considered poor, between 0.40 and 0.54 weak, between 0.55 and 0.69 moderate, between 0.70 and 0.84 good and above 0.84 was considered excellent.^35^ Finally, we recalculated the accuracy after applying the pooling algorithms described above.

## Results

Overall, 17 ChatGPT models existed at the point of the study. Given that GPT-3.5 has been discontinued, we included 16 models in our analysis. Each model was prompted in 10 trials with all 45 cases, resulting in an assessment of 7,200 cases. Both the inter-rater agreement of the coders classifying the model output manually and the inter-rater agreement of the two NLP models were very high (ICC = 0.997 and ICC = 0.996, respectively). After manually assessing and coding disagreements between the NLP models (24/7200 cases), the classification models classified all cases (100%) correctly according to the manual classification ground truth.

On average, o1-mini demonstrated the highest accuracy with 78% of cases solved correctly (331/450, [95% CI 69-78%]), whereas gpt-4.1-nano had the lowest accuracy with 44% of cases solved correctly (196/450, [95% CI 39-48%]). Although all models correctly identified all emergency cases, o1-mini had the highest accuracy in correctly classifying non-emergency cases (100%, 300/300, [95% CI 99-100%] and o4-mini the highest accuracy for self-care cases (22%, 69/130, [95% CI 44-62%]), see Figure 1.

**Figure 1:**
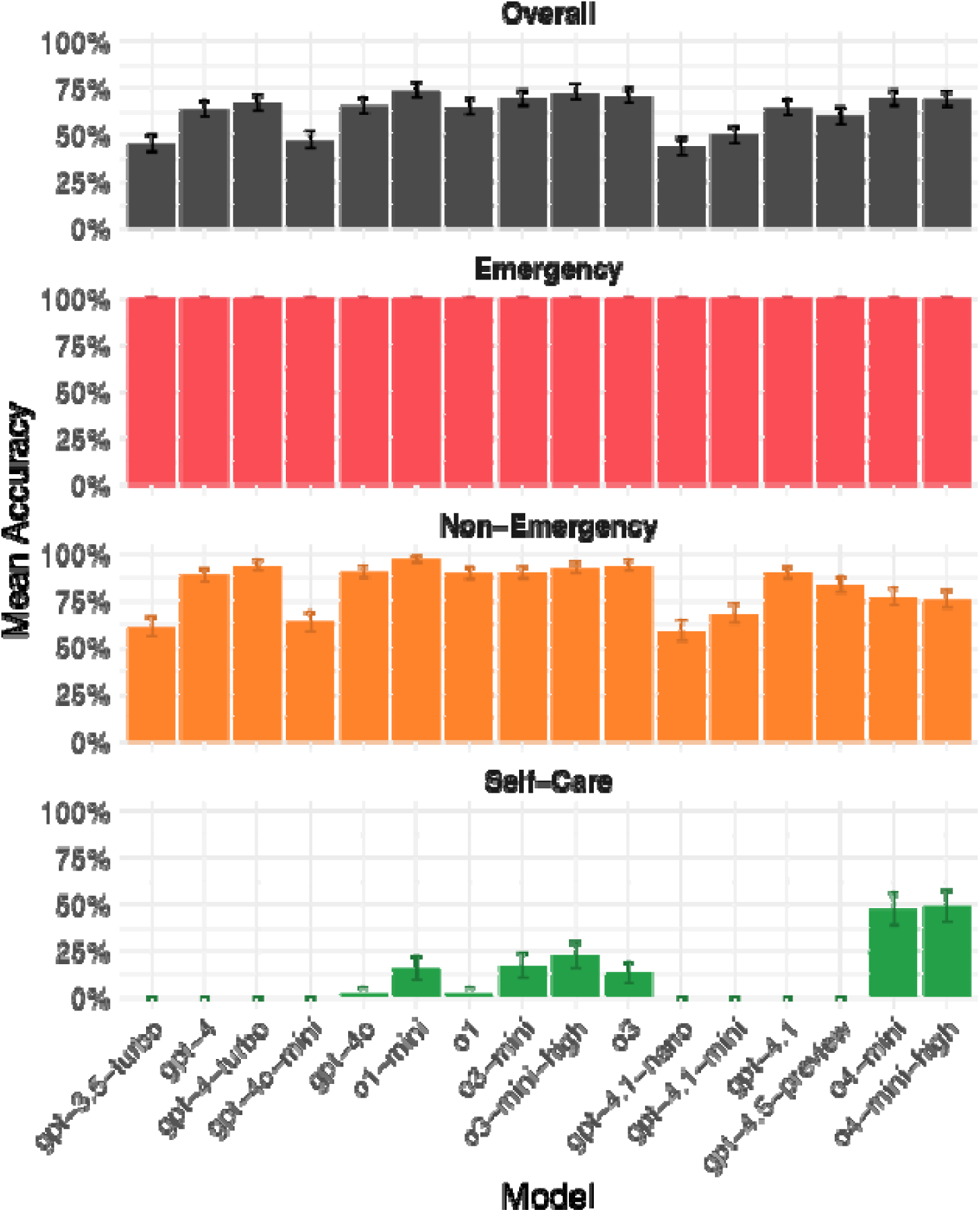
Accuracy of all ChatGPT models sorted by release date (from left to right) across different urgency levels. Emergency accuracy may be unreliable because the vignette set contained only two emergency cases.

All models had the tendency to assign a higher level of urgency than the gold standard solution. This tendency is particularly evident for gpt-3.5-turbo, where all cases were assigned an urgency level that was either correct or higher than necessary. In newer models, the tendency to overtriage decreased.

Specifically, the proportion of overtriage errors among all errors decreased from 100% (247/247, [95% CI 100-100%]) for gpt-3.5-turbo to 69% (96/140, [95% CI 60-76%]) for o4-mini-high, see Figure 2.

**Figure 2:**
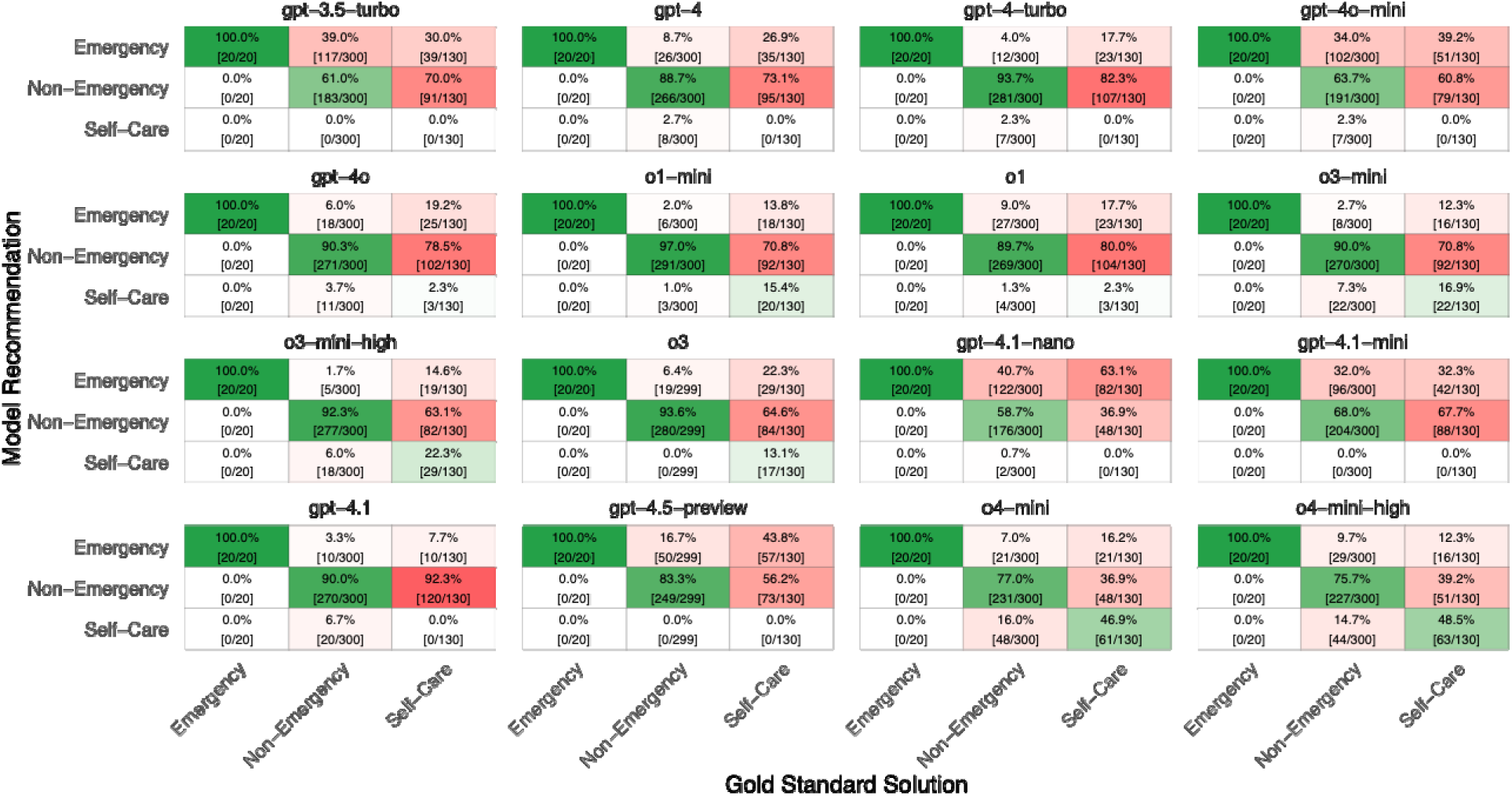
Confusion matrix of urgency levels based on gold standard solutions (x-axis) versus ChatGPT model recommendations (y-axis). Incorrect recommendations are shown in red, whereas correct recommendations are green.

The output for the same case across multiple trials was variable, and this variability tended to increase with newer models. Cohen’s Kappa was moderate to high for all models, see Table 1. Whereas older models had a high proportion of cases that were never solved correctly (49%, 22/45, [95% CI 34-64%] for gpt-3.5-turbo), newer models had a lower proportion (9%, 4/45, [95% CI 2-21%] for o4-mini-high). However, newer models had a higher proportion of cases that received inconsistent advice across multiple trials, that is, that were solved correctly in some trials and incorrectly in other trials (42.2%, 20/45, [95% CI 30-60%] for o4-mini-high and 11%, 5/45, [95% CI 3-24%] for gpt-3.5-turbo), see Figure 3. Notably, only one model (gpt-4.5-preview) was either always or never correct, without any variability.

**Figure 3:**
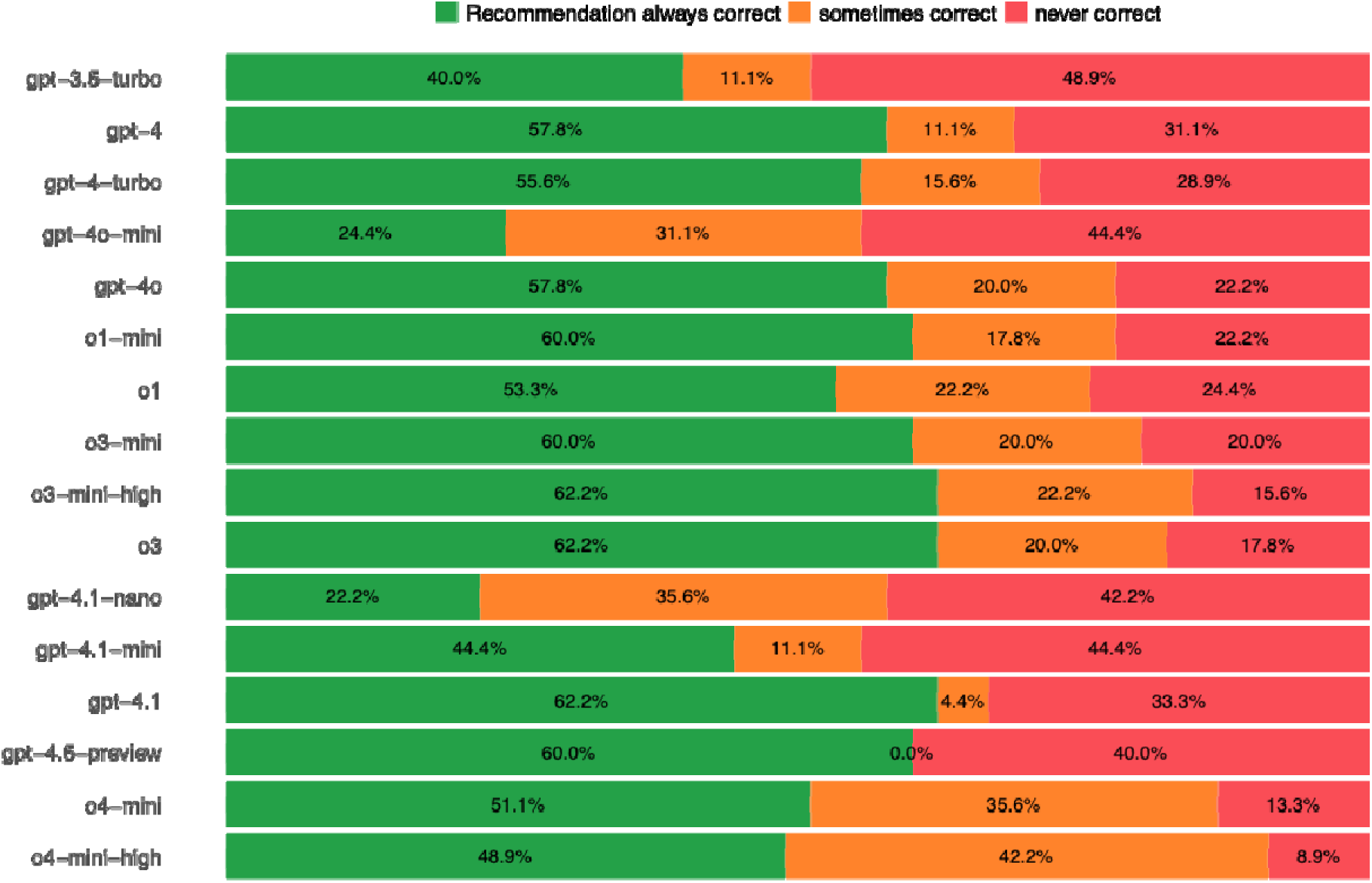
Variability of recommendations per case, for 45 cases and ten trials. The values indicate how often all ten trials yielded the same recommendation that was either correct (green) or incorrect (red) for the same case or whether recommendations differed within ten trials (orange).

**Table 1:** Interrater agreement of all ChatGPT models across ten trials.

| Model | Cohen's Kappa |
| --- | --- |
| gpt-3.5-turbo | 0.84 |
| gpt-4 | 0.81 |
| gpt-4-turbo | 0.68 |
| gpt-4o | 0.71 |
| gpt-4o-mini | 0.67 |
| o1-mini | 0.69 |
| o1-preview | 0.67 |
| o3-mini | 0.71 |
| o3-mini-high | 0.75 |
| o3 | 0.71 |
| gpt-4.1-nano | 0.67 |
| gpt-4.1-mini | 0.82 |
| gpt-4.1 | 0.96 |
| gpt-4.5-preview | 0.97 |
| o4-mini | 0.71 |
| o4-mini-high | 0.70 |

Results of the pooling rules are displayed in Figure 4. Not surprisingly, performance across all models improved the most compared to their overall accuracy by coding a correct solution once the model solved a case correctly at least once across all ten trials (M_all_models_ = +9 percentage points, ranging from 4 to 12 percentage points). The second highest performance improvement across all models was achieved when coding the lowest urgency advice across all trials as the final advice (M_all_models_ = +4 percentage points, ranging from -0.2 to 8 percentage points). Following the majority rule resulted in a decreased performance (-0.4 percentage points, ranging from -2 to 4 percentage points), as well as using the highest urgency advice across all trials (M_all_models_ = -6 percentage points, ranging from -16 to -1 points). Notably, self-care accuracy improved substantially – albeit not all models gave self-care advice – when using the lowest urgency across all trials (M_all_models_ = 19 percentage points, ranging from 8 to 28 percentage points). For example, o4-mini-high could solve 77% (10/13, [95% CI 46-95%]) of self-care cases correctly using this pooling algorithm, whereas taking the mean across all trials resulted in a self-care accuracy of 48% (63/130, [95% CI 15-30%]), see Figure 4.

**Figure 4:**
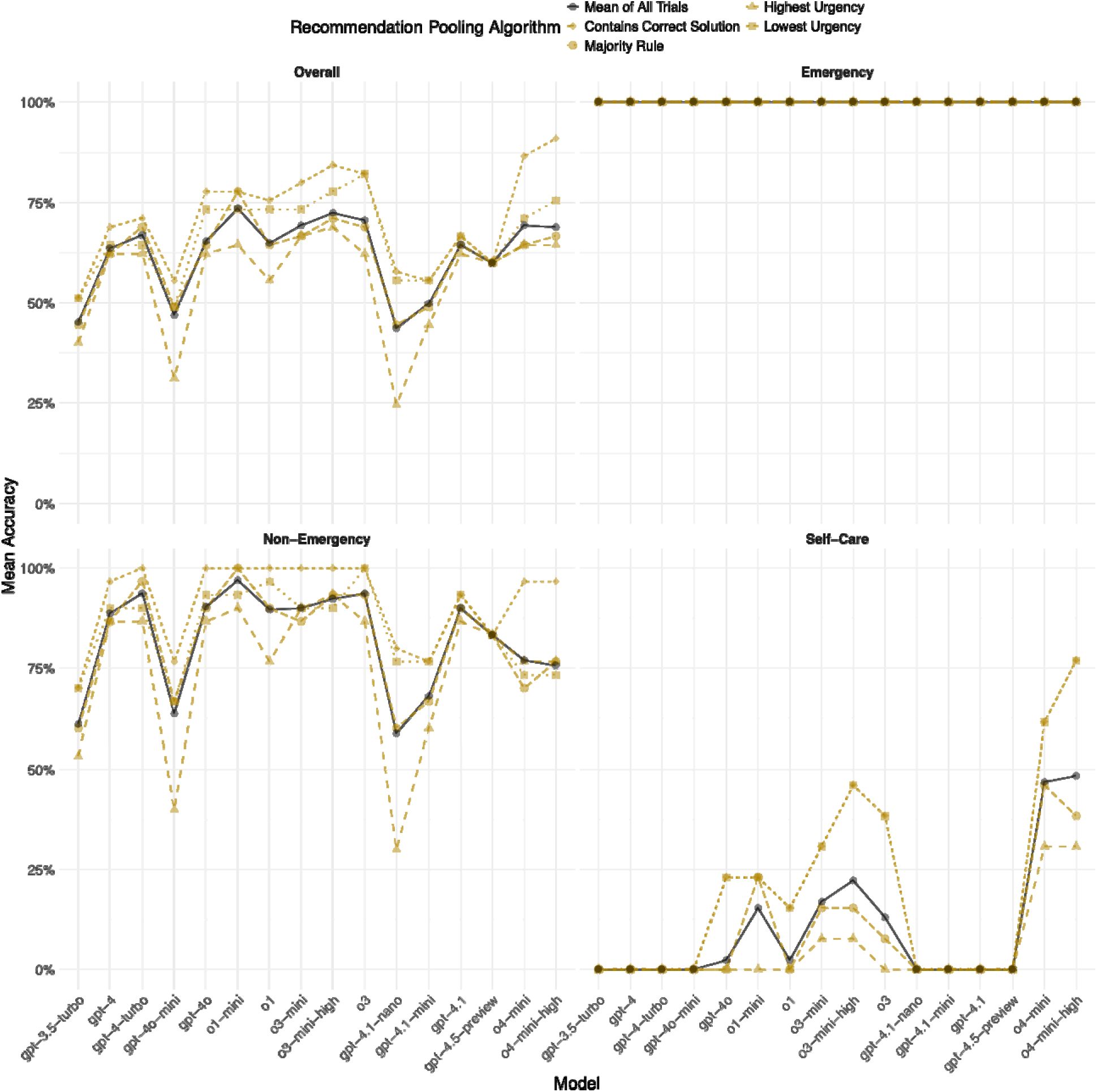
Accuracy of all ChatGPT models when recommendations are pooled according to various algorithms. Emergency accuracy may be unreliable because the vignette set contained only two emergency cases.

## Discussion

In this study, we benchmarked all available ChatGPT models to assess the accuracy of the care-seeking advice they provide using real patient cases. Our results show that average accuracy remains relatively constant across models and does not improve with newer iterations. However, self-care accuracy has improved over time, particularly with the introduction of CoT models and o4-mini. It should be noted, however, that variability in recommendations also increased with newer models. To account for this variability, we explored pooling the recommendations for the same case across multiple trials using different algorithms. Our results indicate that – because the models appear to be risk-averse – using the lowest urgency recommendation may be an effective strategy to increase the accuracy of LLMs. These findings are discussed next.

In general, our accuracy estimates align with those reported in previous studies and tested with various vignette sets. For example, a previous study using the same case vignettes reported an accuracy of 71% for GPT-4, and another study using different case vignettes reported an accuracy of 67%.^23,24^ A third study, which tested both gpt-3.5 and gpt-4 with a third set of case vignettes, reported an accuracy of 59% for gpt-3.5 and 76% for gpt-4, respectively.^7^ In other words, existing estimates match our findings for the same models, even though all studies used different case vignettes, which we interpret as evidence for the validity of the existing data and our results.

Our study extends previous research by including and comparing a larger number of models. Results show that the average accuracy of all ChatGPT models remains relatively stable at approximately 70%, with only the oldest model, gpt-3.5-turbo, and smaller models like gpt-4o-mini, gpt-4.1-nano, and gpt-4.1-mini performing markedly worse. Despite these deviations, we did not observe any significant improvement in average accuracy across newer models. However, we observed that self-care recommendations are more readily provided with the advent of CoT models. In a previous study, Levine et al. hypothesized that the low proportion of self-care recommendations might be due either to a lack of self-care discussions in the online data used to train the models or to a need for common-sense reasoning to arrive at a self-care decision.^16^ Given that our self-care cases were identified online, and because self-care recommendations increased with CoT models – which include a reasoning process before generating recommendations – our findings suggest that increased common-sense reasoning may be the driver. In fact, one explanation for the performance increase in CoT models is that their reasoning approach may circumvent safety measures that previously prevented models from providing self-care advice to minimize potential harm to users.

Despite the increase in self-care recommendations, the accuracy of identifying self-care cases remains far from perfect and currently offers limited decision support for users. An additional factor limiting the usefulness of self-care advice by existing ChatGPT models is the substantial variability in the outputs.

Because self-care is only occasionally recommended across multiple trials, even if it is the correct solution, users are likely to be advised to seek medical care more often than necessary. Given that laypeople generally recognize emergency cases reliably but have problems determining whether self-care is an appropriate option,^24,36^ future models and applications that build on these models should strive to further improve the accuracy of the self-care advice if their goal is to assist in care-seeking decisions.

Our analyses suggest that a potential strategy to further increase (self-care) accuracy could involve prompting the model multiple times and aggregating its recommendations. As demonstrated in our analysis of pooling algorithms, selecting the lowest urgency recommendation across multiple trials may currently be the most promising procedure. This approach improves accuracy in identifying self-care cases while maintaining high accuracy in identifying emergency and non-emergency. Given that laypeople typically do not ask the same question multiple times,^36^ CoT models may apply the pooling algorithm on its own answers to improve the care-seeking advice it provides.

### Practical Implications

This study has several implications for practice and regulation. First, the current and previous findings indicate that currently available ChatGPT models are effective for identifying emergencies.^16,23^ Medical laypeople should therefore use them if they are uncertain whether emergency care is necessary. For cases other than emergencies, most ChatGPT models are currently not reliable tools, in particular if users are unsure whether they require medical care or can rely on self-care and stay at home. Only o4-mini and o4-mini-high outperformed laypeople, although they still correctly identified only half of all self-care cases.^24^ However, prompting these models multiple times and selecting the lowest urgency recommendation can substantially increase self-care accuracy, with only minor reductions in correctly identifying emergency and non-emergency cases. Because some models perform better with self-care cases while others perform better with non-emergency cases, we recommend that users select and use the low-urgency pooling algorithm with o4-mini or o4-mini-high when completely uncertain about the appropriate care level.

Conversely, if users can confidently exclude self-care, they should use other CoT-models such as o1 or o3 as they have shown higher accuracy in this regard than o4-mini and o4-mini-high.

This study also demonstrates that AI technology should be evaluated and regulated according to specific use cases, despite its claims to domain generality. Although ChatGPT models seem to have high utility in some use cases such as text summarization, self-diagnosis, and answering general medical questions, the usefulness of most models in terms of care-seeking advice is limited.^4–9,12,14,15^ In fact, by frequently recommending a higher urgency level than medically necessary, these models may actively contribute to increased and potentially inappropriate healthcare utilization, with the current economic impact of misguided resource utilization estimated at over four billion dollars annually already.^37,38^ This risk is further exacerbated by high variability in outputs and the fact that self-care was rarely advised across multiple trials, even when it was the most appropriate recommendation. Future evaluation methods and regulations should thus explicitly specify the use case AI tools are tested for and should also find ways to deal with variability in LLM output. They should also test multiple models and identify those that perform best in a selected use case. As our findings demonstrate, accuracy varies depending on how multiple outputs for the identical prompt are pooled. If evaluators rely solely on ‘technical accuracy’ (i.e., whether the correct solution was provided at least once), they risk overestimating the model’s real-world accuracy. Instead, to obtain a more accurate estimate of its real-world impact, using the mean of all recommendations may be a more effective approach, as it better reflects the advice (and variability) received by an average user.

### Limitations and Ways Forward

Our study has several limitations. First, the prompt used in this evaluation is artificial, and laypeople are unlikely to phrase their queries in the same way. However, prompting strategies vary widely among users, and the exact wording depends entirely on the individual using it.^36^ We adopted a prompt used in previous studies to ensure cross-study comparability and to reduce potential bias because of input variability of different users.^16,23,24^ Additionally, we used a standardized and validated vignette set and benchmarking methodology to assess the accuracy of all models.^24^ If future studies want to evaluate the impact in real-world scenarios, i.e., how ChatGPT influence users’ decisions, they should consider observational designs or clinical studies involving real patients who generate their own prompts based on their own symptoms.

The second limitation concerns the gold standard solutions to the case vignettes, which were determined by two physicians but represents only an approximation to the ground truth. Establishing the ground truth would require a comprehensive clinical assessment, including lab tests, imaging, and long-term follow-up, to accurately determine the cause of symptoms and thus the most appropriate urgency level.^39^ However, we followed established guidelines in developing the gold standard solution, and our findings align with those of other studies in which urgency ratings were provided by different experts.^16,23^

The last limitation concerns the distribution of urgency levels in our dataset. The case set was developed to reflect natural base rates of all urgency levels, but true medical emergencies among users of care-seeking decision aids in the real world are relatively rare. Consequently, only a small number of emergency cases (2) were included, which may have led to an overestimated accuracy for this urgency level. However, similar studies with a higher number of emergency cases have reported consistently high accuracy in identifying emergencies, suggesting that this limitation does not critically undermine our conclusions.^16,23^

## Conclusion

Our study demonstrates that although the overall accuracy of ChatGPT models in providing care-seeking advice does not improve, these models have become better––but certainly not sufficiently––at identifying cases where self-care is appropriate. Because self-care is the most valuable form of advice for users, the current utility of most ChatGPT models as decision support systems for care-seeking decisions remains limited and only o4-mini and o4-mini-high with an additional advice-pooling algorithm may be useful.

Prompting these models multiple times and selecting the lowest urgency recommendation can increase their accuracy in identifying self-care cases. Because there is currently no one-size-fits-all model––some models perform better with more urgent cases, while others perform better with low-urgency cases––users should carefully select the most appropriate ChatGPT model or another non-LLM application for their specific use case rather than relying on a single general-purpose model.

## Data availability

The data will be published in an open access repositorium upon acceptance.

## Author contributions

MK and MAF conceived of the study. MK developed the procedure. MK and LH collected the data. MK conducted the analyses and data visualization and wrote the first draft of the manuscript. All authors provided critical input and worked on manuscript development.

